# Changing social contact patterns among US workers during the COVID-19 pandemic: April 2020 to December 2021

**DOI:** 10.1101/2022.12.19.22283700

**Authors:** Moses C. Kiti, Obianuju G. Aguolu, Alana Zelaya, Holin Y. Chen, Noureen Ahmed, Jonathan Battross, Carol Y. Liu, Kristin N. Nelson, Samuel M. Jenness, Alessia Melegaro, Faruque Ahmed, Fauzia Malik, Saad B. Omer, Ben A. Lopman

## Abstract

Non-pharmaceutical interventions minimize social contacts, hence the spread of SARS-CoV-2. We quantified two-day contact patterns among USA employees from 2020–2021 during the COVID-19 pandemic. Contacts were defined as face-to-face conversations, involving physical touch or proximity to another individual and were collected using electronic diaries. Mean (standard deviation) contacts reported by 1,456 participants were 2.5 (2.5), 8.2 (7.1), 9.2 (7.1) and 10.1 (9.5) across round 1 (April–June 2020), 2 (November 2020–January 2021), 3 (June–August 2021), and 4 (November–December 2021), respectively. Between round 1 and 2, we report a 3-fold increase in the mean number of contacts reported per participant with no major increases from round 2–4. We modeled SARS-CoV-2 transmission at home, work, and community. The model revealed reduced relative transmission in all settings in round 1. Subsequently, transmission increased at home and in the community but remained very low in work settings. Contact data are important to parameterize models of infection transmission and control.

**Teaser:** Changes in social contact patterns shape disease dynamics at workplaces in the USA.

## Introduction

Over the last two years, estimation of empirical social contact patterns has been reinvigorated following the emergence of severe acute respiratory syndrome-corona virus-2 (SARS-CoV-2), the virus that causes coronavirus disease-19 (COVID-19). Social contact pattern data are critical to understand spread of respiratory pathogens such as SARS-COV-2 and assess the effectiveness of control efforts. Contact studies mainly use self-reported data via contact surveys to quantify “who-contacts-whom”, with typical stratifications by age, setting, and other disease-related attributes (*1, 2*). These patterns vary at multiple geographic scales primarily due to population structure, culture, and socio-economic activities (3, 4). Epidemiologically, workers represent an important population due to potential exposure to respiratory pathogens such as flu and SARS-CoV-2 at work (*5*), increased risk of severe infection with age (*6*), and the potential to transmit infections to household members during lockdowns (*7, 8*). Mathematical models have been widely used to simulate the transmission of SARS-COV-2 and examine the impact of different patterns of social contacts on control (*9*). However, patterns and rates of contacts at workplaces are poorly understood in the US (*10*).

Population-based contact studies conducted during the first year of the COVID-19 pandemic reported significant reductions in contact rates compared to periods before March 2020 (*11*). In the Spring and Summer of 2020, contact rates in North America, Western Europe and Asia dropped to 2–5 contacts per person from 7 to 26 contacts reported during pre-pandemic periods. In March 2020, local, state, and federal authorities in the US recommended non-pharmaceutical interventions (NPIs), including stay-at-home orders and closures of schools and nonessential workplaces, to decrease contact rates aiming to reduce transmission of SARS-COV-2 (*12*). Between April and December 2020, telework accounted for an estimated 50% of paid work hours (*13*), and more than 98% (n=304) of respondents in a survey targeting 3 companies reported ever working from home during the period April through June (*2*). Non-Hispanic Blacks, those aged <45 years, and males, reported higher contact rates and longer duration interactions with other household members compared to other groups [9]. When lockdowns were relaxed in Fall 2020 and Spring 2021, workplace contacts in retail, hospitality and transportation sectors reported a rebound in the number of contacts (*14*), as demonstrated by the drop in the Stringency Index (*15*) (range 0 to 100 depending on how stringent the physical distancing containment measures were). However, the mechanisms and impact of physical distancing interventions on SARS-CoV-2 transmission across time remains poorly understood.

Starting in April 2020, we conducted a cross-sectional study to collect data on social contact patterns among employees in 3 companies in Atlanta, Georgia, USA (*2*). In subsequent rounds, these companies plus 2 others provided data at three additional timepoints up to December 2021. In this report, we describe the changing contact patterns among employees during the ongoing COVID-19 pandemic in the US.

## Results

### Description of study participants

Across four rounds of data collection, 1,456 respondents reported a total of 12,198 contacts. Participation increased modestly from R1 (N=304) to R4 (N=433) with no major fluctuations observed in the proportions across rounds by age, sex, race, and ethnicity. However, only 16 individuals participated in all four rounds. In total, about one third of participants (n = 442) were aged 20–29 years and 5% (n = 80) were 60 years and older. Among all participants, 64% (n = 933) were female. The majority (n = 1,293; 89%) of participants had a bachelor’s degree or higher. The family structure varied from living alone (n = 222; 15%), in a nuclear family (n = 919; 63%), 9% in extended families, with roommates (10%) and the rest in other arrangements. Close to two–thirds of the participants were white (n = 909; 62%) and 7% (n = 95) of Hispanic ethnicity. At the time of the study for each round, >95% of all participants reported ever working from home. In R4, 14% (60/433) of individuals reported ever having COVID-19 confirmed by a test. Out of all participants in R4, 97% (n=420) reported having received any COVID-19 vaccine. A summary of the participants’ characteristics is provided in Table 1.

**Table 1.** Baseline characteristics of study participants. This shows the number of participants across 4 rounds of data collection in five US companies, April 2020 – December 2021. NH refers to non-Hispanic ethnicity.

|  | **Total**<br>**(N (%))** | **Round 1** | **Round 2** | **Round 3** | **Round 4** |
| --- | --- | --- | --- | --- | --- |
|  | **N = 1,456** | **N = 304** | **N = 343** | **N = 376** | **N = 433** |
| **Sex** |  |  |  |  |  |
| Female | 933 (64) | 184 (61) | 227 (66) | 248 (66) | 274 (63) |
| Male | 518 (36) | 116 (38) | 115 (34) | 128 (34) | 159 (37) |
| Not reported | 5 (0) | 4 (1) | 1 (0) | 0 (0) | 0 (0) |
| **Age (years)** |  |  |  |  |  |
| 20–29 | 442 (30) | 90 (30) | 87 (25) | 120 (32) | 145 (33) |
| 30–39 | 413 (28) | 76 (25) | 109 (32) | 104 (28) | 124 (29) |
| 40–49 | 320 (22) | 60 (20) | 86 (25) | 80 (21) | 94 (22) |
| 50–59 | 201 (14) | 49 (16) | 39 (11) | 56 (15) | 57 (13) |
| 60+ | 80 (5) | 29 (10) | 22 (6) | 16 (4) | 13 (3) |
| **Education** |  |  |  |  |  |
| Lower than Bachelors | 162 (11) | 17 (6) | 35 (10) | 51 (14) | 59 (14) |
| Bachelors or higher | 1,293 (89) | 286 (94) | 308 (90) | 325 (86) | 374 (86) |
| **Family structure** |  |  |  |  |  |
| Live alone | 222 (15) | 44 (14) | 43 (13) | 60 (16) | 75 (17) |
| Nuclear | 919 (63) | 173 (57) | 241 (70) | 236 (63) | 269 (62) |
| Extended | 138 (9) | 26 (9) | 27 (8) | 40 (11) | 45 (10) |
| With roommates | 146 (10) | 39 (13) | 28 (8) | 37 (10) | 42 (10) |
| Other | 31 (2) | 22 (7) | 4 (1) | 3 (1) | 2 (0) |
| **Race/ Ethnicity** |  |  |  |  |  |
| Hispanic | 95 (7) | 14 (5) | 20 (6) | 24 (6) | 37 (9) |
| Asian, NH | 281 (20) | 48 (16) | 37 (11) | 75 (20) | 121 (28) |
| Black, NH | 133 (9) | 25 (8) | 30 (9) | 35 (9) | 43 (10) |
| White, NH | 847 (59) | 169 (56) | 240 (70) | 226 (60) | 212 (55) |
| Mixed, NH | 71 (5) | 46 (15) | 12 (4) | 13 (3) | 0 (0) |
| Other, NH | 29 (2) | 2 (1) | 4 (1) | 3 (1) | 20 (5) |
| **Working from home** |  |  |  |  |  |
|  | 1396 (96) | 288 (95) | 329 (96) | 368 (98) | 411 (95) |

### Contact patterns

Across all rounds, the least contacts were reported at the workplace (1,647, 14%), while a third of the contacts were reported at home (4515, 37%) and about half in the community (6036, 49%) (see SI.3). The median (IQR) number of contacts over both days reported in R1, R2, R3 and R4 was 2 (1–4), 7 (4–10), 7 (4–12) and 8 (4–13), respectively (Table 2). The median number of contacts in R2 was 3.5 times higher than R1 and this was sustained to R4. Corresponding mean (standard deviation) values over both days for each round are 2.5 (2.5), 8.2 (7.1), 9.2 (7.1) and 10.1 (9.5), respectively. The increase was consistent across age, sex, and education level. Between R1 and R4, however, we observed a 6–fold and 2.5–fold increase in median number of workplace and community contacts, respectively, whereas no change was reported at the household. We also present a summary of median contacts by setting in SI.4.

**Table 2.**
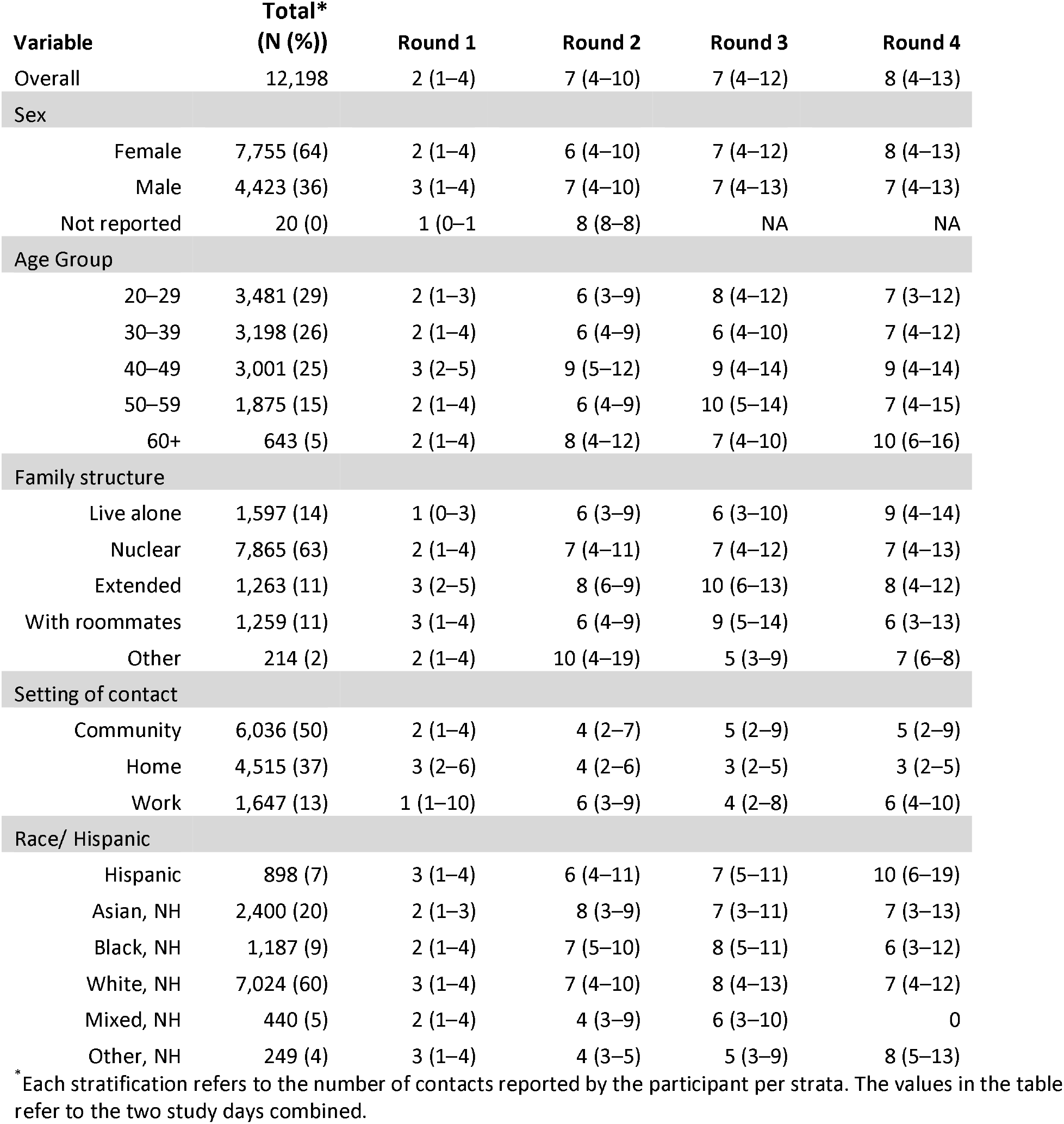
Distribution of contacts. This shows the median and interquartile range (IQR) of contacts reported by participants across four rounds of data collection in five US companies, April 2020 – December 2021. NH refers to non-Hispanic ethnicity.

| Variable | Total*<br>(N (%)) | Round 1 | Round 2 | Round 3 | Round 4 |
| --- | --- | --- | --- | --- | --- |
| Overall | 12,198 | 2 (1–4) | 7 (4–10) | 7 (4–12) | 8 (4–13) |
| Sex |  |  |  |  |  |
| Female | 7,755 (64) | 2 (1–4) | 6 (4–10) | 7 (4–12) | 8 (4–13) |
| Male | 4,423 (36) | 3 (1–4) | 7 (4–10) | 7 (4–13) | 7 (4–13) |
| Not reported | 20 (0) | 1 (0–1) | 8 (8–8) | NA | NA |
| Age Group |  |  |  |  |  |
| 20–29 | 3,481 (29) | 2 (1–3) | 6 (3–9) | 8 (4–12) | 7 (3–12) |
| 30–39 | 3,198 (26) | 2 (1–4) | 6 (4–9) | 6 (4–10) | 7 (4–12) |
| 40–49 | 3,001 (25) | 3 (2–5) | 9 (5–12) | 9 (4–14) | 9 (4–14) |
| 50–59 | 1,875 (15) | 2 (1–4) | 6 (4–9) | 10 (5–14) | 7 (4–15) |
| 60+ | 643 (5) | 2 (1–4) | 8 (4–12) | 7 (4–10) | 10 (6–16) |
| Family structure |  |  |  |  |  |
| Live alone | 1,597 (14) | 1 (0–3) | 6 (3–9) | 6 (3–10) | 9 (4–14) |
| Nuclear | 7,865 (63) | 2 (1–4) | 7 (4–11) | 7 (4–12) | 7 (4–13) |
| Extended | 1,263 (11) | 3 (2–5) | 8 (6–9) | 10 (6–13) | 8 (4–12) |
| With roommates | 1,259 (11) | 3 (1–4) | 6 (4–9) | 9 (5–14) | 6 (3–13) |
| Other | 214 (2) | 2 (1–4) | 10 (4–19) | 5 (3–9) | 7 (6–8) |
| Setting of contact |  |  |  |  |  |
| Community | 6,036 (50) | 2 (1–4) | 4 (2–7) | 5 (2–9) | 5 (2–9) |
| Home | 4,515 (37) | 3 (2–6) | 4 (2–6) | 3 (2–5) | 3 (2–5) |
| Work | 1,647 (13) | 1 (1–10) | 6 (3–9) | 4 (2–8) | 6 (4–10) |
| Race/ Hispanic |  |  |  |  |  |
| Hispanic | 898 (7) | 3 (1–4) | 6 (4–11) | 7 (5–11) | 10 (6–19) |
| Asian, NH | 2,400 (20) | 2 (1–3) | 8 (3–9) | 7 (3–11) | 7 (3–13) |
| Black, NH | 1,187 (9) | 2 (1–4) | 7 (5–10) | 8 (5–11) | 6 (3–12) |
| White, NH | 7,024 (60) | 3 (1–4) | 7 (4–10) | 8 (4–13) | 7 (4–12) |
| Mixed, NH | 440 (5) | 2 (1–4) | 4 (3–9) | 6 (3–10) | 0 |
| Other, NH | 249 (4) | 3 (1–4) | 4 (3–5) | 5 (3–9) | 8 (5–13) |

\* Each stratification refers to the number of contacts reported by the participant per strata. The values in the table refer to the two study days combined.

**Table 3.** Baseline characteristics of study participants. This shows the number of participants across 4 rounds of data collection in five US companies, April 2020 – December 2021.

**Table 4.** Distribution of contacts. This shows the median and interquartile range (IQR) of contacts reported by participants across four rounds of data collection in five US companies, April 2020 – December 2021.

A 9-fold increase in median number of contacts was also noted in individuals who lived alone, from a median of 1 (IQR 0–3) to 9 (4–14) in R1 to R4, respectively, as shown in Table 2.

### Contact matrices across rounds

Figure 2 shows changing age–specific contact patterns across the four rounds on a graduated scale in employees of five US companies.

**Figure 1.**
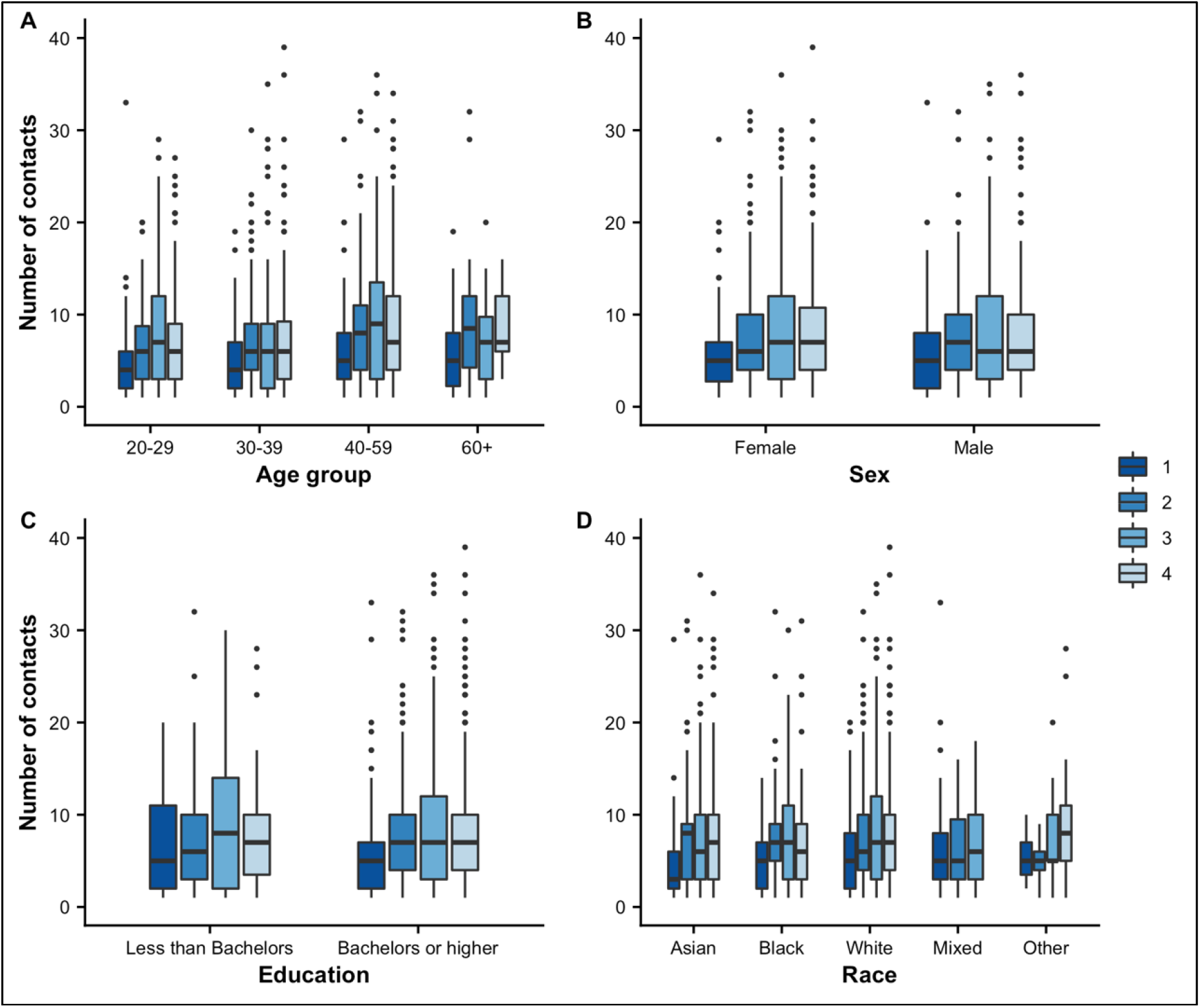
Median (IQR) contacts over two days by various participant attributes in five US companies, April 2020 – December 2021. Panels (**A**), (**B**), (**C**) and (**D**) show the distribution of reported contact by age group, sex, education level and race for R1–R4.

**Figure 2.**
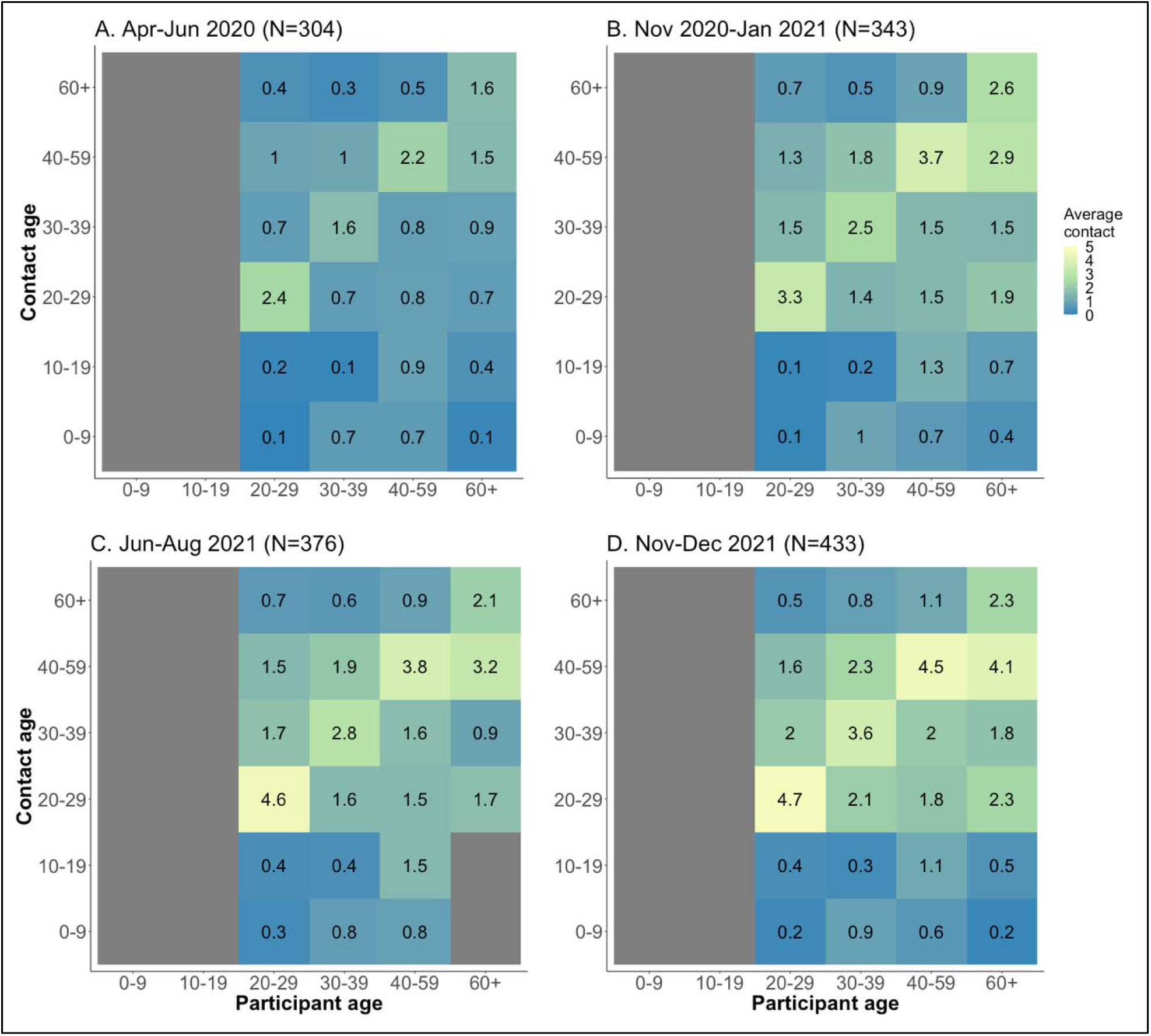
Contact matrices showing the mean number of contacts over two days for each round in employees of five US companies. Panel (**A**) shows contacts in R1 (Apr–Jun 2020), (**B**) shows R2 (Nov 2020–Jan 2021), (**C**) shows R3 (Jun–Aug 2021), and (**D**) shows R4 (Nov–Dec 2021). The gray column on ages 0–19 years indicates no contacts reported since all participants are employees aged >20 years. The gray bar between age 60+ and 0–19 years in panel C indicates that no contacts were reported.

Across all rounds, we observe two key characteristics. The first is the presence of the prominent diagonal (assortative contacts), signifying a higher number of contacts between people of the same age. The number of assortative contacts increased subtly through the rounds. The second observation is the presence of interactions between 30–39 and 40–59–year-olds with children and young adults aged 0–19 years old. These contacts remain relatively stable across rounds. Lastly, in later rounds, we observed more contacts off the diagonal, indicating that contacts become less assortative as individuals started interacting more across different ages.

### Difference of contact patterns between rounds

In Figure 3, we show the net difference in the age specific average number of contacts occurring only at the workplace between consecutive rounds (panel A–C) and the first and last rounds (R1– R4, panel D). The highest net positive change was observed in ages 30–39 years and the least to no change was observed in the oldest group (60+ years). An increase in the average number of contacts was observed from rounds 1–2 and 2–3, while some ages (20–29 and 40+ years) showed net decreases.

**Figure 3.**
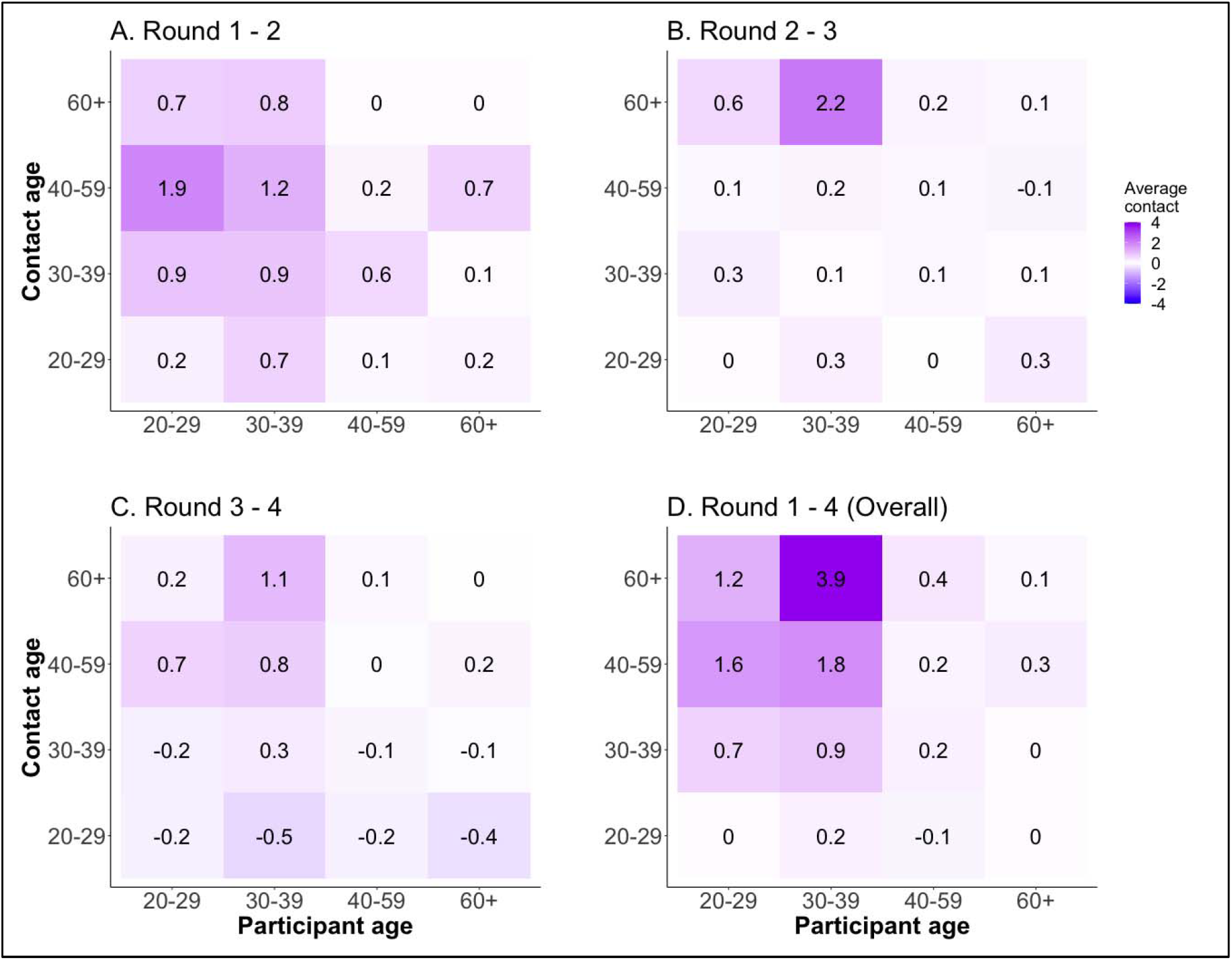
Matrices of difference in contacts between rounds in employees of five US companies. The panels show differences between R1–R2 (A), R2–R3 (B), R3–R4 (C), and R1–R4 (D), respectively.

### Contact patterns by setting

We also assess the mixing patterns by age in Figure 4 separately for work (panel A–D), home (E–H) and community (I–L) across the four rounds. We observed differences in the number and structure of contacts across settings and rounds. Work contacts increase marginally across rounds and occur across all age groups. Home contacts displayed distinct assortative mixing patterns that increased marginally in R2 compared to R1 and do not change thereafter. We also observed the presence of intergenerational contacts between parents (30–59 years) and children (0–19 years). Community contacts displayed the highest net increase from R1–R4 with both assortative contacts and contacts between people of different ages. At home and in the community, contacts were generally high among young adults aged 20–29 years.

**Figure 4.**
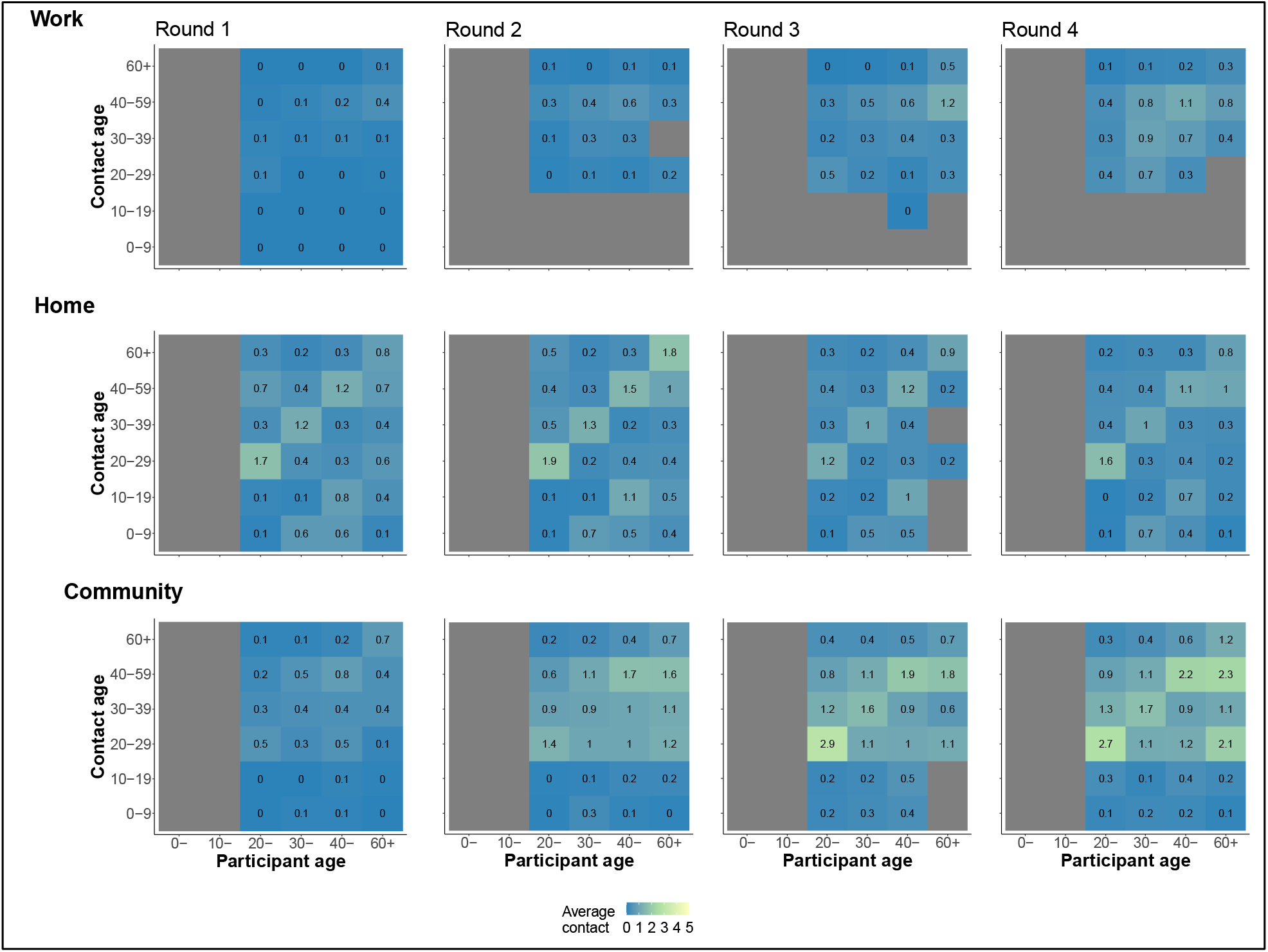
Matrices of evolution of contacts occurring exclusively at work, home, and community across rounds in employees of five US companies. The top panel shows contacts at work, middle panel shows contacts at home, and bottom panel contacts in the community across study rounds.

### Impact of changing contacts on SARS-CoV-2 transmission potential

We estimate the impact of changing social contact on SARS-CoV-2 transmission. In round 1, reductions in contact relative to pre-pandemic periods suppressed the relative transmissibility to substantially below 1 at work and in the community but had a smaller effect at home. Increases in age-specific contacts between rounds 1 and 4 led to an increase in the relative transmissibility with varying effects across settings (Figure 5). We estimated relative transmission to increase more at community settings such as stores, parks, and gyms than at work settings across study rounds. For all rounds, we observed that the relative transmissibility at work remained below 1. On the other hand, relative transmissibility in community settings rose after round 1 but stayed similar between rounds 2 through 4 and remained below one.

**Figure 5.**
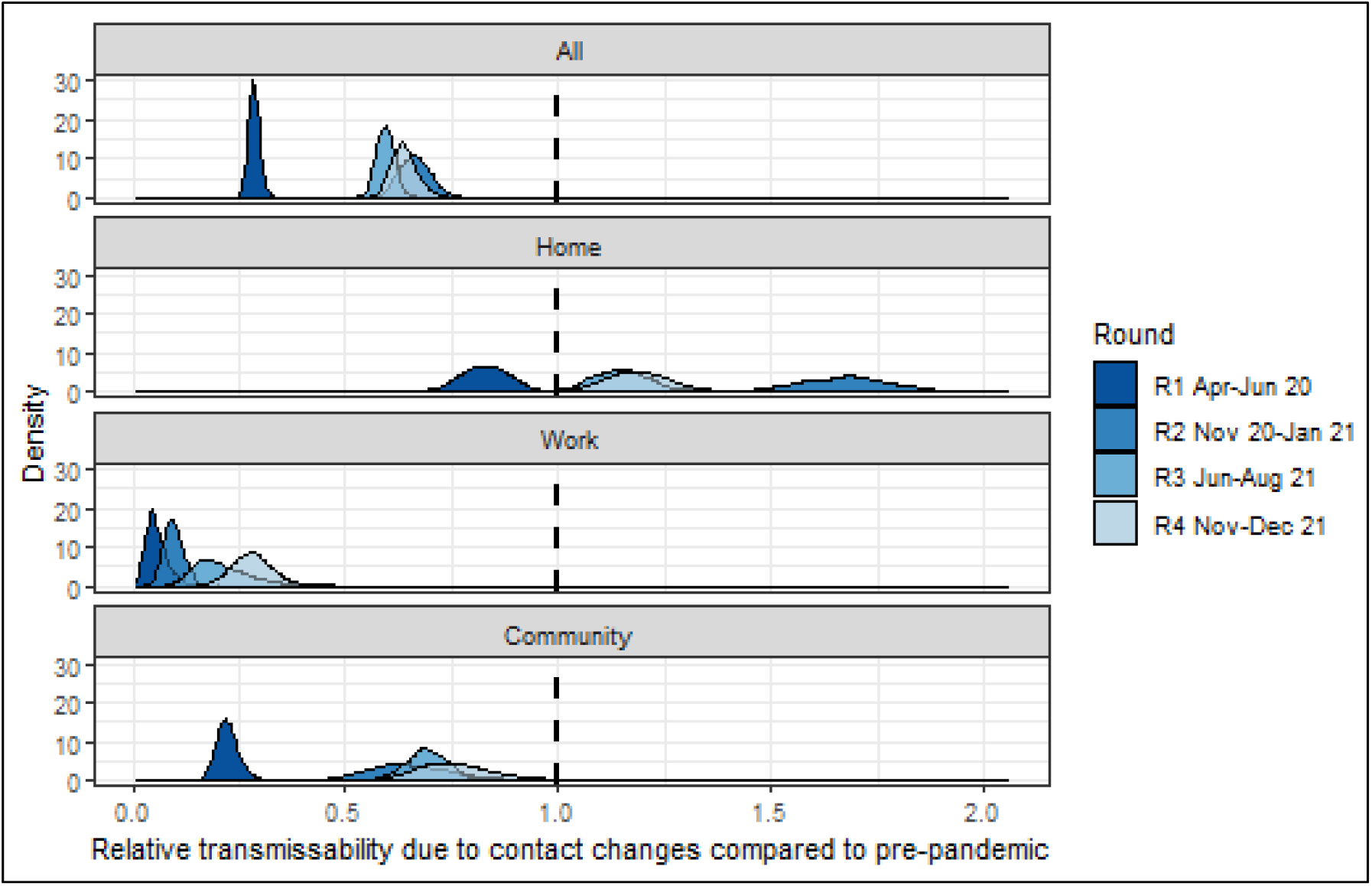
Changes in transmissibility due to changes in age–specific contact patterns alone in employees of five US companies. The relative transmissibility is inferred by comparing rounds 1– 4 of age-specific contact patterns to projected baseline age-specific matrices for the US (*16*). On the x-axis, 1.0 denotes no change in relative transmissibility, values <1.0 denote reduced transmissibility and values >1.0 denote increased transmissibility. The y-axis denotes the probability density.

**Figure 6.** Median (IQR) contacts over two days by various participant attributes in five US companies, April 2020 – December 2021. Panels A, B, C and D show the distribution of reported contact by age group, sex, education level and race for R1–R4.

**Figure 7.** Contact matrices showing the mean number of contacts over two days for each round in employees of five US companies. Panel A shows contacts in R1 (Apr–Jun 2020), B shows R2 (Nov 2020–Jan 2021), C shows R3 (Jun–Aug 2021), and D shows R4 (Nov–Dec 2021). The gray column on ages 0–19 years indicates no contacts reported since all participants are employees aged >20 years. The gray bar between age 60+ and 0–19 years in panel C indicates that no contacts were reported.

**Figure 8.** Matrices of difference in contacts between rounds in employees of five US companies. The panels show differences between R1–R2 (A), R2–R3 (B), R3–R4 (C), and R1– R4 (D), respectively.

**Figure 9.** Matrices of evolution of contacts occurring exclusively at work, home, and community across rounds in employees of five US companies. The top panel shows contacts at work, middle panel shows contacts at home, and bottom panel contacts in the community across study rounds.

**Figure 10.** Changes in transmissibility due to changes in age–specific contact patterns alone in employees of five US companies. The relative transmissibility is inferred by comparing rounds 1–4 of age-specific contact patterns to projected baseline age-specific matrices for the US (*16*). On the x-axis, 1.0 denotes no change in relative transmissibility, values <1.0 denote reduced transmissibility and values >1.0 denote increased transmissibility. The y-axis denotes the probability density.

## Discussion

This study quantified social contact patterns among workers in selected companies in the US at multiple timepoints during the COVID-19 pandemic period from April 2020 to December 2021. Participants in our study reported a large increase in the median number of contacts between April 2020–June 2020 and November 2020–January 2021 across all age groups and in both workplace and community (non-household) settings. Contacts remained high after January 2021. We leveraged these data to estimate the impact of changing social contact patterns on SARS-CoV-2 transmission. In our model, we observed reduced transmissibility of SARS-COV-2 compared to transmission that would have occurred in the absence of physical distancing policies. The extent of reduction differed by setting of contact (home, school, or community). Our new findings suggest that workers reported substantial increases in the rates of contact during the study period which were an independent driver of SARS-CoV-2 transmission.

Overall, contacts were very low between April–June 2020 (median = 2) coinciding with the stringent containment measures at that time. Employees from all companies we surveyed were working from home and interactions were largely limited to family members or roommates. Contacts peaked in round 2 of data collection from November 2020 to January 2021 (median 8) with the highest percentage increase noted at work. However, the reported average number of contacts remained lower than that compared to pre-pandemic periods captured by the European POLYMOD study with mean contacts ranging between 8 (Germany) to 20 (Italy) (*3*). Similarly, by Spring 2021, multiple studies in the US (*17, 18*) reported high number of contacts reported at work. Community contacts also increased and became more heterogeneous across time as workers interacted with a wider pool of individuals. However, despite the relaxation of physical distancing policies, the average number of contacts reported per person did not rise above pre-pandemic levels.

We observed reduced transmission potential in the workplace when more stringent containment measures were in place (April–June 2020) compared to later periods with rollback (round 2–4, from November 2020). Our model suggests increased transmissibility in the home (transmission rate above 1 relative to pre-pandemic periods) and marginally in the community (remaining less than 1) after restrictions were rolled back. Transmissibility at work increased marginally despite significant increases in the number of contacts at work. Increased mobility outside the home and corresponding increases in heterogeneous number of contacts at work compared to earlier pandemic periods have also been observed due to easing of restrictions (*17, 19*). Despite bans on gathering in US states including Georgia, we expected that contacts would have been higher than reported in this study after November 2020 due to increased mobility and home visits, potentially resulting in the infection surges observed after the 2020 Thanksgiving and Christmas holiday periods (*20*). Our results, highlighting low contact numbers during early phase of the pandemic, are consistent with previous studies in the US (*17, 21*), UK (*19*) and China (*22*). Studies that collect data on changing contact patterns over time and in various settings remains important at this stage in the pandemic. With the persistence of individuals hesitant to get vaccinated (*23*) and the emergence of more transmissible variants (*24*), and limited understanding of the extent of SARS-CoV-2 immunity (*25*), there remains the need to use empirical social contact data and mathematical models to better inform workplace infection prevention policies such as frequency of testing, work-from-home, and mandated adequate protection for those who cannot telework. This research has some limitations. First, this was an opt-in survey administered online to employees of five companies, thus subject to selection bias. This was different from some other surveys that have used existing population panels (*18*) or conducted random sampling of the population (*17, 19*). We were unable to get the exact number of individuals and demographic composition to whom the survey links was sent so we could not compare the demographic composition of our respondents to the company workforce. Our respondents were highly educated, majority White individuals working in private companies. Thus, we cannot claim representativeness of the study sample to the US workforce. However, some of the findings have been shown in other studies, which suggest that the current sample does not appear to differ in a meaningful way from a general sample of people in the USA. To encourage higher survey uptake, we offered a $40.00 gift card upon completion of each survey and held meetings with employees to inform them of study progress and explain the importance of our studies. Lastly, we assumed that the change in transmissibility was due to changes in contact patterns only despite the implementation of other public health interventions including mask wearing and availability of vaccines from round 2 (Nov–Dec 2020). In our estimates for relative transmissibility, we assume a fully susceptible population and that transmissibility of SARS-CoV-2 is invariable by age. Moreover, as no empirical data from the US were available prior to the pandemic, we used published estimates inferred from European contact structure (*16*) which may be less reflective of pre-pandemic contacts in the US. Despite these limitations, our findings on reduced transmission were similar to previous modeling studies.

In conclusion, we present a unique study that observed changing contact patterns among members of a specific sector of the U.S. workforce during the ongoing COVID-19 pandemic. We found that the transmission of SARS-CoV-2 was dependent on setting-specific contact patterns. While the social contact patterns were used to understand changes in human behavior during the SARS-CoV-2 outbreak and its impact on SARS-CoV-2 transmission, these data are also relevant for other endemic pathogens such as influenza that are transmitted through close contacts.

## Materials and Methods

### Experimental design

The objective of this study was to characterize the patterns of social contact and mixing in non-healthcare workplace settings in select large companies in the United States using standardized social contact diaries. This was an online cross-sectional study recruiting participants from five private companies based in Georgia, US. These companies include workers falling under the “educational services”, “management occupations”, “business and financial operations occupations”, “computer and mathematical occupations” and “life physical and social science occupations” sectors as defined by the US Bureau of Labor Statistics (*26*). Between April 2020 and December 2021, we conducted four rounds of data collection: April–June 2020 (Round 1, abbreviated as R1), November 2020–January 2021 (R2), June–August 2021 (R3), and November–December 2021 (R4). Individuals could participate in multiple rounds. R1 represents a transition period of non-pharmaceutical interventions leading to the Stringency Index dropping from highs of 70 in April to <60 in June (*15*). On 1^st^ May 2020, mandatory stay-at-home orders were lifted for persons at low risk of infection in the state of Georgia (*27*) where most of our participants resided and 98% had reported working from home (*2*). R2 occurred during the large SARS-COV-2 winter wave in 2020 when schools were closed, and masking was mandatory in selected spaces (*15*). R3 and R4 occurred when most of the containment measures had been rolled back, and the latter round occurred during the Omicron surge in the winter of 2021 (*24*). During R3 and R4, vaccinations were widely available in the US (*28*).

### Data collection

Recruitment procedures were as described previously for R1 (*2*). Individuals voluntarily opted into the study. On enrolment, we collected data on participant demographics (age, sex, education, race, job role, family size and composition, current residence, and work setting) and company details (name, office size, teleworking schedule).

One day following enrollment, each participant received a weblink to complete a survey to report the number of individuals with whom they had a contact with over two continuous workdays (Monday to Friday). All contacts irrespective of setting were reported. We defined a contact as either proximate (no conversation and no physical contact but within 6 feet of another person for more than 20 seconds, e.g., sitting next to someone in public transport or standing in line), conversational (a two-way conversation with three or more words exchanged in the physical presence of another person), or physical (directly touching someone (skin-to-skin contact) or the clothes they are wearing, intentionally or unintentionally, including a handshake, fist bump, elbow bump, foot bump, hug, and kiss). The 20–second duration was selected to capture the fastest social interactions between individuals in a social setting (*29*). For each contact, participants recorded their age in years (0–9, 10–19, 20–29, 30–39, 40–59, 60+), sex (male, female), relationship to participant, setting of contact, and participation in perceived higher-risk activities such as attending school, work, indoor/ outdoor gatherings, gym, going to restaurants, living in a nursing home, or air travel. Setting of contact was categorized as home, work, and community, whereby community represented all other areas apart from home and work. All other definitions remain the same as reported in R1 (*2*). The full questionnaire is available in Supplementary Information 1 (SI.1).

### Statistical analyses

All analyses were performed with R v4.1.2. All code and data are available on SOCRATES (*30*), an online platform for sharing social contact data.

#### Descriptive statistics

We described characteristics of participants by age (20–29, 30–39, 40–49, 50–59 and 60+ years old), sex (male, female), race (Asian, Black, White, Mixed or Other), ethnicity (Hispanic or not), and family structure. Family structure was categorized as living alone, nuclear family (combination of respondent, spouse, and children), extended family (nuclear family plus relatives), or living with unrelated roommates. All companies circulated the survey link to their employees living and working in the USA.

#### Average contacts

We calculated the median number of contacts per person and their associated interquartile ranges (IQR). We report contacts by age groups, sex, race, ethnicity, family structure and setting of contact. Unless otherwise stated, all analyses in the main text include contact made cumulatively over both survey days; single day contacts are reported in SI.2.

#### Contact matrices by age

We divided the age group-specific number of contacts by the number of participants in that age group. Contact matrices were stratified by round and setting of contact. We used four age groups for the participants (20–29, 30–39, 40–59, 60+ years) consistent with R1 data and six age groups for the contacts (0–9, 10–19, 20–29, 30–39, 40–59, 60+ years) (*2*).

#### Impact of social contacts on SARS-COV-2 transmission

We estimated the impact of changing social contact patterns on SARS-CoV-2 transmission by comparing age-specific contact patterns for each round to synthetic pre-pandemic contact rates (henceforth called “baseline”) for the US as derived from the POLYMOD study (3, 16). We used a published method (31) to derive the relative changes in transmission due to changes in social contacts from the ratio of dominant eigenvalues of the age-specific contact matrix. This approach assumed that infectiousness and susceptibility did not vary by age group. We also assumed that schools remained closed during our study data collection periods and thus did not account for contacts that may have occurred at school. Since children <18 years of age did not participate in our study, we generated square matrices by imputing child-child and child-adult contacts. Imputation was done by using the ratio between the dominant eigenvalues of matrices from each study round to the baseline matrix. Bootstrapping was done using the socialmixr R package (*32*).

### Ethics statement

Ethical approval was given by Yale University (IRB# 2000026906). All participants signed an electronic informed consent form. Participants received a $40 gift card upon completion and submission of the questionnaire. All data were de-identified before analysis.

## Data Availability

All data produced in the present study are available upon reasonable request to the authors.

## Acknowledgments

We thank the employees of the five companies that participated in this study.

## Author contributions

Funding acquisition: SBO, BL, FA

Conceptualization: SBO, BL

Validation: MCK, AZ, FM

Methodology: MCK, OGA, AZ, NA, CYL, KNN, SJ, AM, FA, FM, BL, SBO

Investigation: MCK, OGA

Formal analysis: MCK, JB, HC, CYL

Visualization: MCK, JB, HC, CYL

Supervision: MCK, OGA, AZ, NA, HC

Writing—original draft: MCK, OGA

Writing—review & editing: MCK, OGA, AZ, NA, KNN, SJ, AM, FA, FM, BL, SBO

Data curation: MCK, HC, JB

Project administration: OGA, AZ, NA, FA

## Disclaimer

The findings and conclusions in this report are those of the authors and do not necessarily represent the official position of the Centers for Disease Control and Prevention.

## Data and materials availability

All data and code are available via our GitHub repository.

## Supplementary Materials

