## Supplementary materials for "Changing social contact patterns among US workers during the COVID-19 pandemic: April 2020 to December 2021"

Supplementary Text

Enrolment form

| Q1 | Please enter your company email address. **(To participate in the survey and to receive your gift card, you must use your official company email address).** |  |
| --- | --- | --- |
| Q2 | Please enter your first and last name: |  |
| Q3 | We would like to be able to send you text messages on your mobile phone with information about study and reminders to complete the survey. Your phone number will not be shared with anyone else and messages will focus on the study.    Please provide your preferred mobile phone number. |  |
| Q4 | Please enter your age. |  |
| Q5 | Which of the following best describes you? | [1] Female  [2] Male  [3] Non-binary |
| Q6 | What is the highest level of education you have completed? | [1] Less than high school degree  [2] High school graduate (high school diploma or equivalent including GED)  [3] Some college but no degree  [4] Associate degree in college (2-year)  [5] Bachelor's degree in college (4-year)  [6] Master's degree  [7] Doctoral degree or Professional degree (e.g. PhD, JD, MD) |
| Q7 | Are you Hispanic, or Latinx or Spanish origin? Select one. | [1] No, not of Hispanic, Latinx, or Spanish origin  [2] Yes, Mexican, Mexican American, Chicano  [3] Yes, Puerto Rican  [4] Yes, Cuban  [5] Yes, another Hispanic, LatinX or Spanish origin |
| Q8 | Which of the following races do you consider yourself to be? | [1] White or Caucasian  [2] Black or African American  [3] American Indian or Alaska Native  [4] Asian  [5] Native Hawaiian or Pacific Islander  [6] Mixed or Multiple Races  [7] Other, please specify |
| Q9 | What is your marital status? | [1] Single  [2] Cohabiting or living with a partner  [3] Married  [4] Separated  [5] Divorced  [6] Widowed |
| Q10 | In which state do you currently reside? |  |
| Q11 | In which state do you currently work? |  |
| Q12 | Please enter the zip code where you currently work. |  |
| Q13 | Who do you live with? Please select all that apply. | [1] I live alone  [2] Spouse/significant other [3] Roommates  [4] Parents  [5] Children  [6] Siblings  [7] Other, please specify |
| Q14 | Including yourself, how many people are currently living or staying at your home?  **INCLUDE:** Everyone who has been living or staying at your home for more than 2 months/ Anyone staying with you who does not have another place to stay, even if they have been with you for 2 months or less. **DO NOT INCLUDE:** Anyone who has been living somewhere else for more than 2 months (i.e., a college student living away or someone in the Armed Forces on deployment) |  |
| Q15 | Which company do you currently work for? |  |
| Q16 | What is your job title? | [1] Individual contributor  [2] Manager  [3] Leader (looks after a region or business area)  [4] Executive/C-Suite  [5] Consultant  [6] Accountant  [7] Human Resources  [8] Landscape Architect  [9] Architect (various levels) (14)  [10] Urban Designer  [11] Interior Designer  [12] Principal  [13] Associate Principal  [14] Marketing  [15] Other, please specify: |
| Q17 | Which of the following best describes the department you work in? | [1] CEO/President  [2] Consulting  [3[ Customer Service/Customer Support  [4] Design (Architect, Urban, Interior)  [5] Education (Administration, Teacher, etc.)  [6] Engineering  [7] Facilities/Office Management  [8] Finance/Accounting  [9] Human Resources  [10] IT  [11] Legal  [12] Medical Plannimg  [13] Marketing/Advertising  [14] Operations/Management  [15 Public Relations  [16] Research & Development  [17] Sales  [18] Supply Chain/Warehouse |
| Q18 | How many employees currently work in your office on a typical day? Do not include anyone teleworking.  We define office as the physical location for work where you interact with a group (clients and/or colleagues) at least twice a week. ***If there are multiple locations that apply, use the location that is most frequent.*** | [1] I work fully remote  [2] 1-4  [3] 5-9  [4] 10-19  [5] 20-49  [6] 50 or more |
| Q19 | Does this office share a building with any other companies or businesses, including shops and restaurants? | [1] Yes  [2] No |
| Q20 | Do you ever work from home/telecommute/telework?  **Telecommute/telework** is defined as work from a location outside of your designated place of work. It may consist of working from home, from a public space outside of your workplace, etc. | [1] Yes  [2] No |
| Q21 | How often did you work from home/telecommute/telework **before** the COVID-19 pandemic? | [1] Never  [2] Less than once a month  [3] Once a month  [4] Multiple times a month  [5] Multiple times a week  [6] Once a week  [7] Every day/Always |
| Q22 | How often do you **currently** work from home/telecommute/telework? | [1] Never  [2] Less than once a month  [3] Once a month  [4] Multiple times a month  [5] Once a week  [6] Multiple times a week  [7] Every day/Always |
| Q23 | Since the pandemic, has your office asked employees to return to in-person work? | [1] Yes, fully in-person required  [2] Yes, fully in-person encouraged not mandatory  [3] Yes, hybrid  [4] No, fully remote |
| Q24 | From where do you primarily work when you telecommute/telework? | [1] Home  [2] Coffee shop  [3] Co-working space  [4] Restaurant  [5] Other, please specify: |
| Q25 | What form of transit do you use to get to this telecommute/telework location? Please select all that apply. | [1] None  [2] Walking or biking  [3] Driving yourself  [4] Carpool with spouse/ friends/ coworkers  [5] Rideshare services  [6] Public transportation  Other, please specify |
| Q26 | Since the pandemic, has your office asked employees to return to in-person work? | [1] Yes, fully in-person required  [2] Yes, fully in-person encouraged not mandatory  [3] Yes, hybrid  [4] No, fully remote |
| Q27 | Have you had COVID-19 (Coronavirus Disease-19)? | [1] Yes, and it was confirmed by a diagnostic test  [2] Yes, but it was not confirmed by a diagnostic test  [3] No  [4] I don't know |
| Q28 | Are you currently ill with COVID-19 or any other infection? | [1] Yes  [2] No  [3] Prefer not to answer |
| Q29 | Have you been vaccinated against COVID-19? | [1] Yes  [2] No  [3] Prefer not to answer |
|  | The following questions pertain to the Flu, please only answer the following questions in regard to the **Flu**. |  |
| Q30 | Q39 Did you get a flu shot this flu season (August 2021 to March 2022)? | [1] Yes  [2] No  [3] I don't know |
| Q31 | How often do you get your flu shot? | [1] Every year  [2] Have before, but not every year  [3] Never |
| Q32 | When was your last flu shot? | [1] After August 2021  [2] July 2019 to August 2020  [3] July 2018 to August 2019  [4] July 2017 to August 2018  [5] Before August 2018 |
| Q33 | Next year, we will collect additional data on social interaction at the workplace using both “Radio Frequency Identification (RFID) proximity sensors”, or sensors for short, and contact diary. We will ask each participant to wear a sensor while working in the office.      These sensors when worn by you and fellow staff will collect data on time, duration, and proximity of contact between you and other participants. The data collected is anonymous. The sensor does not record audio or video. Further instructions will be provided in full before we start this phase of data collection.      Would you consider participating in this sensor component of this study next year? | [1] Yes  [2] No |
| Q34 | Any comments or questions about the sensor component of this study: |  |
| Q35 | May we contact you to participate in similar studies in the future? | [1] Yes  [2] No |

CorporateMix diary

| Q1 | Please enter your company email address.  **(To participate in the survey and to receive your gift card, you must use your official company email address).** |  |
| --- | --- | --- |
| Q2 | **Introduction**  Within the next week, we would like you to record in the diary **every** person that you have **contact with** over two consecutive **work** days. You will complete the online contact diary at the end of the second day. Please find attached a Memory Aid that you can use to keep track of your contacts throughout the two days. Each day should cover a 24-hour period. The data from this diary will help us learn how people interact in the community, which will in turn help us understand how some respiratory diseases such as COVID-19 and influenza may spread from one person to another, and how we can control the spread. You may print the memory aid for use to keep track of your contacts throughout the day. At the end of the second day, you will complete an online contact diary using the information you recorded in the memory aid.  **For this research, a contact is defined as:**  **Physical contact:** directly touching someone (skin-to-skin contact), or the clothes they are wearing intentionally or unintentionally (for example, a handshake, fist bump, elbow bump, foot bump, hug, kiss, etc.)  **Non-physical contact**: a two-way conversation with three or more words exchanged in the physical presence (within 2 m or approximately 6 ft of each other) but with no physical contact.  **Direct proximity:** being within 6 feet of your contact for 20 seconds or more with neither conversation nor physical contact.  If you contact the same person more than once during the day (e.g., first encounter: 5 mins; second encounter:30 mins), only record them once, but enter the total combined time you spent with that person (e.g., 35 mins in total).  After you finish recording your contacts in the diary, please double check the diary entries to make sure you have not left out any people with whom you had a contact. The order in which you write down your contacts is not important. We suggest using a chronological order, starting with the first person you had a contact with during each of the days, and then adding everyone else as you went through your daily activities. | |
| Q3 | What company do you currently work for? |  |
| Q4 | Please enter the date (i.e., mm/dd/yyyy) of the **first** day you are reporting contacts for. |  |
| Q5 | Please enter the date (i.e., mm/dd/yyyy) on the second day you are reporting contacts for. |  |
| Q6 | Did you work in-office or did you telework your first day **on** | [1] In-office (defined as a physical location where you interact with a group at least twice a week)  [2] Telework (working from a location outside of your designated place of work. Teleworking may consist of working from home, from a public space outside of your workplace, etc.)  [3] Mix of both |
| Q7 | Did you work in-office, or did you telework your **second day**? | [1] In-office (defined as a physical location where you interact with a group at least twice a week)  [2] Telework (working from a location outside of your designated place of work. Teleworking may consist of working from home, from a public space outside of your workplace, etc.)  [3] Mix of both |
| Q8 | **On either day**, were you in any large group situations where you had contacts but could not identify everyone individually (e.g., meetings, grocery store, hospitals, public performances, movie theaters, etc.)? | [1] Yes, day 1 only  [2] Yes, day 2 only  [3] Yes, both days  [4] No contact with large groups |
| Q9 | How many people did you come into contact (as defined in the previous question) with your **first day**? |  |
| Q10 | How many people did you come into contact with on the second day **only?**  If you came into contact with someone on the first day or both days, **DO NOT COUNT THEM HERE.** |  |
| Q11 | Please enter a unique identifier for the person you are currently identifying.  This identifier can be anything that helps you identify this person in case you have contact on day 2, such as nicknames (i.e., coworker 1, cashier at Starbucks, Nurse 1, first name, etc.). |  |
| Q12 | What is this person's gender? | [1] Female  [2] Male  [3] Non-binary  [4] I don't know |
| Q13 | Please choose the estimated age range of this person. | [1] Less than 1 year old  [2] 1 to 9 years old  [3] 10 to 19 years old  [4] 20 to 29 years old  [5] 30 to 39 years old  [6] 40 to 49 years old  [7] 50 to 59 years old  [8] 60 to 69 years old  [9] 70 to 79 years old  [10] 80 years and older |
| Q14 | What is your relationship with this person? | [1] A household member (e.g., family members such as spouse, child, parent, or non-family member such as roommate)2  [2] Relative, not a household member  [3] Friend/acquaintance  [4] Colleague  [5] Do not know this person personally |
| Q15 | How often do you have contact with this person, in general? | [1] Daily or almost daily  [2] About 1 to 2 times per week  [3] About 1 to 2 times per month  [4] Less than once per month  [5] I don't have contact with this person generally |
| Q16 | Over the past 14 days, please indicate whether this person has or has not participated in the below activities to the best of your knowledge | |
|  | \| Yes \| No \| I don't know; attended in-person class (e.g., daycare, K-12 school, college) \| \| \|; attended large gatherings outdoors (e.g., sporting events or concerts, amusement parks etc) \| \| \|; worked in a healthcare setting \| \| \|; worked in a shared office at least once a week \| \| \|; participated in high contact sports (e.g., basketball, football) \| \| \|; attended in-person gathering indoors at least once a week (e.g., church, choir, parties, meetings) \| \| \|; worked out in a gym at least once a week \| \| \|; had dinner inside a restaurant/bar or visited movie theater at least once a week \| \| \|; used public transit \| \| \|; traveled by air \| \| \|; Lived in a group home or nursing home \| \| \| | |
| Q17 | Did you make physical contact and/or speak with this person? Please select all that apply. | [1] Non-physical contact  [2] Physical contact  [3] Direct proximity |
| Q18 | Where did you come into contact with this person? | [1] Home  [2] Another person's home  [3] Gym  [4] Healthcare setting (e.g., hospital, clinic, nursing home)  [5] Place of worship  [6] Playground  [7] School/College  [8] Store/Mall  [9] Transport/Hub  [10] Street  [11] Work  [12[ Salon/Barber shop  [13] Restaurant/eatery  [14] Other, please specify |
| Q19 | What was the total time you spent with this person during the entire day (including overnight)? | [1] Less than 5 minutes  [2] Between 5 to 15 minutes  [3] Between 15 minutes to 1 hour  [4] Between 1 hour to 4 hours  [5] More than 4 hours |
| Q20 | Was this person properly wearing a mask (i.e., covers nose and mouth) during your contact with them? | [1] Yes, for the entire encounter  [2] Yes, during parts of encounter  [3] No, mask was not worn at all during encounter  [4] I don't recall |
| Q21 | Did you also have contact with [person listed in Q21] on your second day? | [1] Yes  [2] No  [3] I don't recall |

Fig. S1. Distribution of total number of contacts over two days by setting for Round 1 to Round 4 in employees of five US companies.

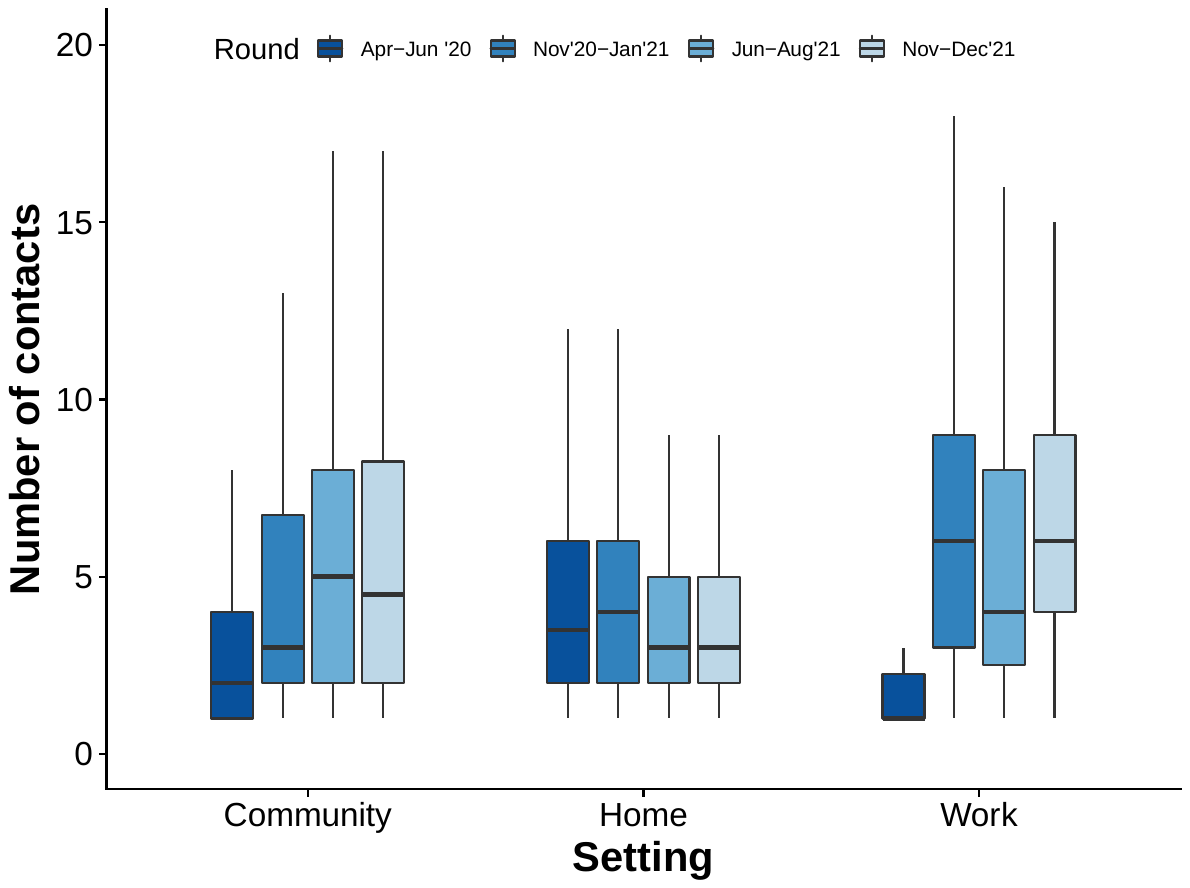

Table S1.

Distribution of participant characteristics and the median and interquartile range (IQR) of contacts reported on the first day only by employees of five US companies.

|  | **Round 1** | **Round 2** | **Round 3** | **Round 4** |
| --- | --- | --- | --- | --- |
| Sex |  |  |  |  |
| Women | 2 (1–4) | 2 (2–6) | 3 (2–6) | 2 (2–6) |
| Men | 3 (1–4) | 2 (2–6) | 2 (2–6) | 2 (2–6) |
| Not reported | 0 (0–1) | 1 (1–1) | NA | NA |
| Age Group |  |  |  |  |
| 20–29 | 2 (1–3) | 2 (2–4) | 3 (2–6) | 2 (2–4) |
| 30–39 | 2 (1–3) | 2 (1–4) | 2 (1–4) | 3 (2–6) |
| 40–59 | 3 (2–5) | 4 (2–6) | 4 (2–6) | 4 (2–6) |
| 50–59 | 2 (2–4) | 4 (2–6) | 2 (1–2) | 2 (2–5) |
| 60+ | 4 (2–4) | 2 (2–6) | 2 (1–2) | 2 (2–5) |
| Family structure | |  |  |  |
| Alone | 0 (0–2) | 1 (1–2) | 2 (1–4) | 2 (1–4) |
| Nuclear | 2 (1–4) | 2 (2–6) | 3 (2–6) | 3 (2–6) |
| Extended | 3 (2–6) | 4 (3–6) | 4 (2–6) | 4 (2–6) |
| Roommates | 3 (2–5) | 2 (2–4) | 2 (2–4) | 3 (2–5) |
| Other | 3 (2–4) | 4 (1–6) | 2 (1–3) | 4 (4–4) |
| Setting of contact | |  |  |  |
| Community | 3 (2–5) | 4 (2–8) | 4 (2.5–8) | 4 (2–7) |
| Home | 2 (1–4) | 4 (2–6) | 3 (2–6) | 2 (2–4) |
| Work | 5 (4–8) | 5 (3–6) | 6 (4–11) | 6 (4–11) |
| Race |  |  |  |  |
| Black | 2 (2–4) | 2 (2–5) | 2 (1–5) | 3 (1–4) |
| White | 2 (1–4) | 2 (2–6) | 3 (2–6) | 3 (2–6) |
| Asian | 3 (1–4) | 2 (1–6) | 3 (2–6) | 2 (2–6) |
| Mixed | 3 (2–4) | 2 (1–4) | 2 (1–2) | 3 (2–5) |
| Other | 3 (1–4) | 4 (2–4) | 2 (2–4) | NA |
| Hispanic |  |  |  |  |
| No | 2 (1–4) | 2 (2–6) | 3 (2–6) | 2 (2–6) |
| Yes | 3 (2–5) | 3 (2–6) | 3 (1–4) | 2 (2–6) |

Table S2.

Number of contacts reported across age, sex, setting, and type of contact in employees of five US companies, April 2020 – December 2021.

|  | **Total**  **(N (%))** | **Round 1** | **Round 2** | **Round 3** | **Round 4** |
| --- | --- | --- | --- | --- | --- |
|  | **N = 12,198** | **N = 1,548** | **N = 2,814** | **N = 3,444** | **N = 4,392** |
| Age of contact |  |  |  |  |  |
| 0–9 | 799 (7) | 139 (9) | 205 (7) | 226 (7) | 229 (5) |
| 10–19 | 885 (7) | 130 (8) | 207 (7) | 291 (8) | 257 (6) |
| 20–29 | 3,229 (26) | 379 (24) | 663 (24) | 943 (27) | 1,244 (28) |
| 30–39 | 2,713 (22) | 302 (20) | 621 (22) | 741 (22) | 1,049 (24) |
| 40–59 | 3,460 (28) | 444 (29) | 830 (29) | 941 (27) | 1,245 (28) |
| 60+ | 1,112 (9) | 154 (10) | 288 (10) | 302 (9) | 368 (8) |
| Sex of contact |  |  |  |  |  |
| Female | 6,143 (50) | 754 (49) | 1,336 (47) | 1,830 (53) | 2,223 (51) |
| Male | 5,823 (48) | 793 (51) | 1,432 (51) | 1,526 (44) | 2,072 (47) |
| Not reported | 232 (2) | 1 (0) | 46 (2) | 88 (3) | 97 (2) |
| Setting of contact |  |  |  |  |  |
| Community | 6,036 (49) | 456 (29) | 1,327 (47) | 1,947 (57) | 2,306 (53) |
| Home | 4,515 (37) | 996 (64) | 1,206 (43) | 1,076 (31) | 1,237 (28) |
| Work | 1,647 (14) | 96 (6) | 281 (10) | 421 (12) | 849 (19) |
| Type of contact |  |  |  |  |  |
| Conversation | 5,872 (48) | 845 (55) | 1,470 (52) | 1,790 (52) | 1,767 (40) |
| Physical | 4,417 (36) | 703 (45) | 1,019 (36) | 1,041 (30) | 1,654 (38) |
| Proximity | 1,909 (16) | – | 325 (12) | 613 (18) | 971 (22) |
